# Patient satisfaction in the use of non-pharmacological therapies in the management of postoperative pain: a study in a tertiary hospital, Ghana

**DOI:** 10.1101/2022.04.28.22274426

**Authors:** Priscilla Felicia Tano, Felix Apiribu, Emile Kouakou Tano, Ramatu Agambire, Isaac Boateng, Victoria Sefah

## Abstract

**Background:** Non-pharmacological therapies are the non-medicinal methods used in postoperative pain (POP) management by health care practitioners such as massage, positioning, breathing exercise, music therapy, and distraction. It has been discovered that therapies that are non-pharmacological for the relief of pain are effective with minimal side effects. Studies have also revealed that non-pharmacological interventions in the management of pain lessen or alter pain perception by decreasing intensity and intensifying the tolerance of pain. This study, therefore, seeks to determine how frequently non-pharmacological methods (NPMs) are used in the management of POP and the influence of these NPMs on patient satisfaction

**Methods:** This study was a descriptive cross-sectional design. A quantitative approach was utilized with a structured questionnaire to obtain responses from patients from October to December 2019. The study was conducted in the female and male surgical wards of Komfo Anokye Teaching Hospital (KATH) in Kumasi, Ghana. 138 patients within the first 72 hours post-operative period who consented and fulfilled the inclusion criteria were enrolled in the study. A convenience sampling method was used to collect the data. Inferential and descriptive statistics were used in analyzing the data.

**Results:** The majority of the patients, 52.2% responded to using some form of (NPMs) for pain relief. The most widely used non-pharmacological method of pain relief was walking representing 79.2%, followed by relaxation 5 (6.8%), distraction 4 (5.6%), listening to music 3 (4.2%), deep breathing 2 (2.8%), and meditation 1 (1.4%). The male participants were observed to use more NPMs of pain relief as compared to the females. Most of the patients (51.4%) who used NPMs for their pain relief reported having their pain not relieved. A majority (90.6%) of the participants desired for more pain treatment. The type of surgery the patients underwent had a statistically significant negative correlation with the use of NPMs of pain relief (Spearman Correlation Coefficien**t** = -0.233, p-value <0.05). A higher percentage (71%) of participants were highly satisfied with the overall pain treatment received.

**Conclusion:** Non-pharmacological management of POP have been proven to be efficient, inexpensive, and have little or no side effect on patients. There should be continuous education on non-pharmacological management of POP to nurses and other healthcare professionals to help promote the efficiency of intervening in postoperative pain with these methods.

## Introduction

Pain treatment is considered a human right as treating pain seeks to gain velocity in WHO policy with this awareness [1]. Surgeries have played a pivotal role in managing and treating complex health challenges in patients requiring the process [2]. However, patients who go through surgical procedures, either minor or major, experience acute POP [3].

Globally the prevalence of POP differs among studies. For instance, the American Pain Society suggests a prevalence of more than 80% with 75% reporting moderate to severe pain [4]. Studies in Sub-Saharan Africa reveal that POP has been undertreated and the burden of POP ranges from 40 to 95% [5,6]. In Ghana, patients who suffer inadequate POP is about 70% [7].

Uncontrolled POP is characterized by longer hospital stays, prolonged post-operative care, more readmission rates, and reduced patient satisfaction [8]. After both major and minor surgeries, the majority of patients have acute POP [9,10]. Study reveals that, POP treatment is not easy to effectively attain because pain is subjective, so finding the type of surgery can aid in achieving successful POP management [11].

Studies suggest that the treatment of POP is still unsatisfactory [1,5,12] even though pharmacological methods are in use for the treatment of pain [13].

Interventions that are not pharmacological are viewed to be therapies that do not include taking medications or other substances that are active [14,15]. More precisely, non-pharmacological therapies are the non-medicinal methods used in postoperative pain management by health care practitioners such as massage, positioning, breathing exercise, music therapy, and distraction. It has been discovered that therapies that are not pharmacological for the relief of pain are effective with minimized side effects and complications [6].

Studies have shown that health care practitioners’ attitude and lack of knowledge on the different types of non-pharmacological pain management makes the efficacious use of these measures questionable [6,16]. Previous studies also reveal that non-medicine interventions in the management of pain lessen or alter pain perception by decreasing intensity and intensifying the tolerance of pain, decreasing pain-related stress, changing pain behavior, reinforcing coping skills, and giving patient and relatives the feeling of control over the pain [17,18,19,20]

Non-pharmacological therapies can complement pharmacological therapies and present another treatment in post-operative pain management but such interventions are under-utilized [14,21]. Using NPMs aids in the reduction of the intake of opioids and its possibly damaging physiological and psychological reactions to the pain [19].

When NPMs are employed as adjuvant treatment, they decrease the anxiety of the patient and increase patient satisfaction with POP management. Several studies on surgical procedures reported that these techniques led to a decreased analgesic use post-operatively, or a reduced pain intensity on Day 2 or 3 of post-operation [22].

NPMs of pain relief can be classified as;

1. Physical activities such as deep breathing, walking, or light to moderate sportive activities.
2. Psychological/spiritual interventions like meditation or relaxation, distraction such as listening to music or talking to people, praying, watching TV, stress management, visualization, hypnosis, and imagery.
3. Inactively used physical methods such as heat / cold packs, massaging, physiotherapy, transcutaneous electrical nerve stimulation (TENS), chiropractic, acupressure, and acupuncture.

Clarity on the understanding of the use of NPMs in our sub-region and the effect on POP management as well as the patient satisfaction with the use of these therapies will be mentioned, as the aim of this study was to understand how frequent the types of NPMs are used, socio-demographics in?uence with the satisfaction of the use of NPMs in the management of POP, if NPMs used, is related to patients’ wish for more pain treatment and the influence of NPMs on patient satisfaction.

## Methodology

### STUDY DESIGN AND STUDY SETTING

This research is part of a larger study of POP management in a tertiary hospital in Ghana where a descriptive cross-sectional design was utilized. A quantitative approach was employed utilizing a structured questionnaire to obtain responses from participants between October 2019 and December 2019. The study was conducted in the female and male surgical wards of Komfo Anokye Teaching Hospital (KATH) in Kumasi, Ghana.

### STUDY PARTICIPANTS, SAMPLING TECHNIQUE, AND SAMPLE SIZE

Post-operative clients within the first 72 hours post-operative period who consented were allowed to participate in the study. Surgical patients who met the inclusion criteria were contacted at the general surgical wards at KATH. The study participants were 138 who were selected using the convenience sampling technique.

### QUESTIONNAIRE

A structured questionnaire created and named International Pain Outcome Questionnaire (IPO-Q) by Pain Out [23] was employed for the collection of data. The IPO-Q is a validated questionnaire that is built on the American Pain Society questionnaire [24]. It uses 11-point (0– 10) NRS elements and binary elements. Two items centering on the duration pain was severe and pain treatment received utilizing percentage scales [24,25]. The questionnaire also comprises a question concerning pain relief experienced by patients and another on patients wanting more pain treatment.

The questionnaire had a section on the use of NPMs which comprised the use of non-medicine treatment to relieve pain and also whether the use of NPMs helped reduce the pain. Also, how often clinicians encourage the use of NPMs as well as the type of NPM used. Studies that employed IPO-Q have shown an acceptable Cronbach’s alpha of 0.85–0.88 88 (26,24,25]

In addition to patient-reported pain outcomes, socio-demographic data (gender, age, marital status, religion, educational level), as well as the type of surgery, duration of surgery, type of anesthesia, type of analgesics given, was obtained through the structured questionnaire.

### DATA ANALYSIS

Inferential statistics, as well as descriptive analysis, were used in the analysis of data which was done using SPSS version 25. In analyzing categorical variables, frequencies and percentages were used.

Pearson’s correlation was employed to determine the correlation strength between the variables. The relationships between demographic variables and NPM used were analyzed by Pearson’s correlation and Spearman Correlation. All tests were conducted at a level of significance of P<0.05.

## Results

### 1. How frequent are the types of NPM used?

Regarding the use of NPMs of pain relief, 72 (52.2%) of the sample population of patients responded to use some form of non-medicinal methods for pain relief whereas 66 (47.8%) resorted only to the use of pharmacological or medicinal methods (Fig 1)

**Figure 1:**
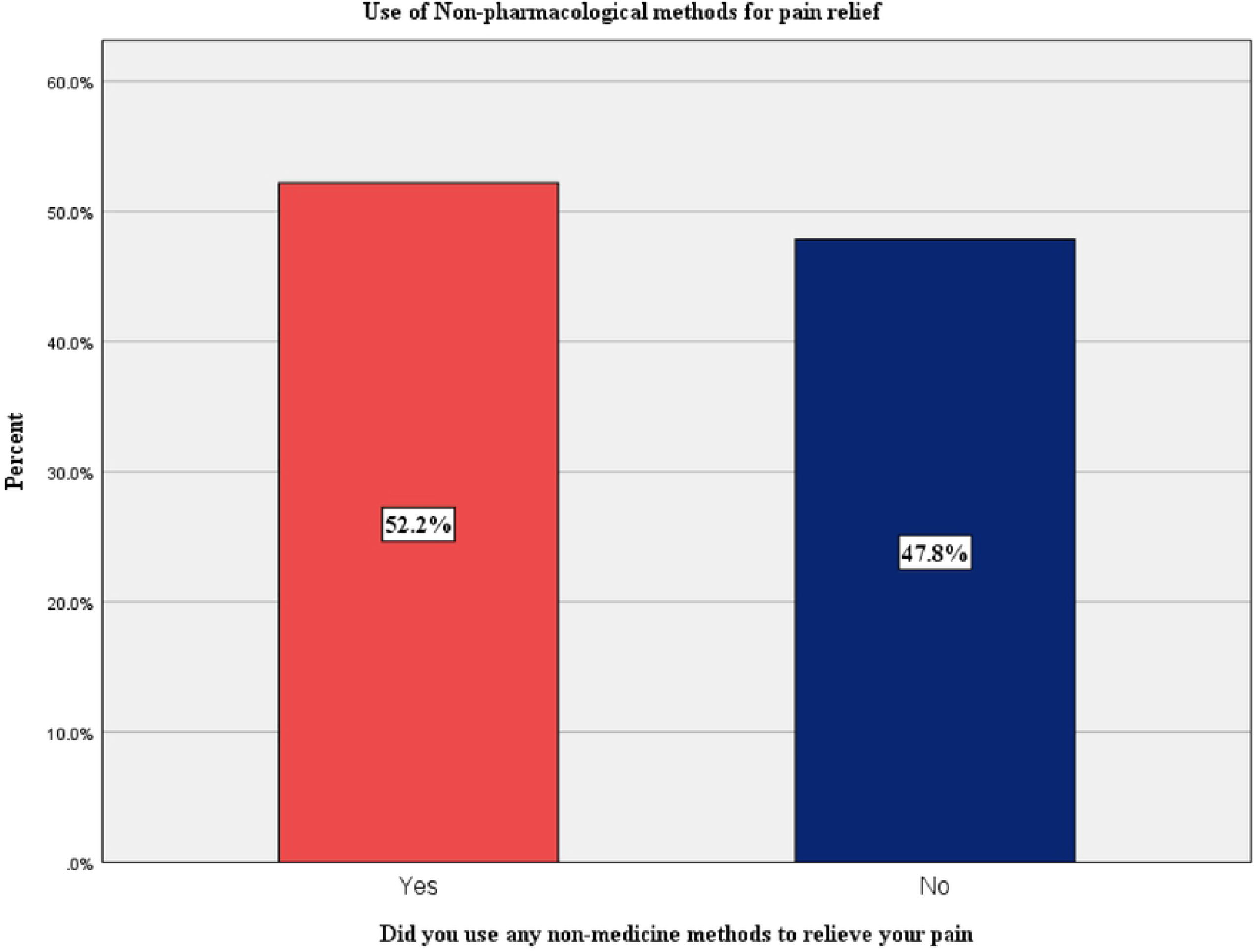
Did you use any non-pharmacological methods to relieve your pain.

From Table 1, the most widely used NPM of pain relief was walking 57 (79.2%), followed by relaxation 5 (6.8%), distraction 4 (5.6%), listening to music 3 (4.2%), deep breathing 2 (2.8%), and meditation 1 (1.4%). The male participants were observed to use more NPMs of pain relief as compared to the females. There was no significant association of use of NPMs between females and males (p-value > 0.05) (Table 1)

**Table 1:** Types of NPMs used for pain relief by gender distribution.

| Non-pharmacological method | Male (n) | Female (n) | Total (n) |
| --- | --- | --- | --- |
| Walking | 28 | 29 | 57 |
| Deep breathing | 2 | 0 | 2 |
| Listening to music | 2 | 1 | 3 |
| Distraction | 2 | 2 | 4 |
| Relaxation | 2 | 3 | 5 |
| Meditation | 1 | 0 | 1 |
| Non-use of NPM | 44 | 22 | 66 |
| Total | 81 | 57 | 138 |

p-value (Cramer's V) = 0.328, Pearson chi-square = 0.328

Moreover, majority of the participants 71 (51.4%) were encouraged by Nurses to use NPMs for their pain relief (Table 2)

**Table 2:** How often did a Nurse encourage you to use NPMs?

| Variable | Frequency (n) | Percent (%) |
| --- | --- | --- |
| Never | 71 | 51.4 |
| Sometimes | 67 | 48.6 |
| Total | 138 | 100.0 |

### 2. Does socio-demographics influence the satisfaction of the use of NPMs in the management of POP?

Meanwhile, a substantial majority of the patients (51.4%) who used NPMs for their pain reliefs reported to have their pain not relieved whereas only 20 (14.5%) had their pain reduced with the use of NPMs (Table 3).

**Table 3:** Were the NPMs helpful in reducing pain?

| Variable | Frequency (n) | Percent (%) |
| --- | --- | --- |
| Yes | 20 | 14.5 |
| No | 71 | 51.4 |
| Total | 91 | 65.9 |

Among the socio-demographic characteristics of the participants, only the type of surgery the patients underwent had a statistically significant negative correlation with the use of NPMs of pain relief (Table 4).

**Table 4:** Socio-Demographic influence on the satisfaction of the use of NPMs in the management of POP.

| Variable | Spearman Correlation Coefficient | p-value |
| --- | --- | --- |
| Gender | 0.077 | 0.468 |
| Age | 0.125 | 0.239 |
| Level of Education | -0.02 | 0.838 |
| Religion | 0.011 | 0.914 |
| Type of Surgery | -0.233* | 0.026 |
| Type of anesthesia | -0.199 | 0.058 |
| Duration of Surgery | 0.022 | 0.838 |
| Prescribed post-operative Analgesia | 0.065 | 0.539 |

\*. Correlation is significant at the 0.05 level (2-tailed).

### 3. Is NPM use related to patients’ desire for more pain treatment?

A substantial majority 125 (90.6%) of the participants in this present study desired for more pain treatment whereas only 13 (9.4%) did not wish for more pain treatment (Table 5).

**Table 5:** Would you have liked more pain treatment than you received?

| Variable | Frequency (n) | Percent (%) |
| --- | --- | --- |
| Yes | 125 | 90.6 |
| No | 13 | 9.4 |
| Total | 138 | 100.0 |

No statistically significant association was identified between the use of NPMs of pain relief and patient desire for more pain treatment (p-value > 0.05) (Table 6)

**Table 6:**
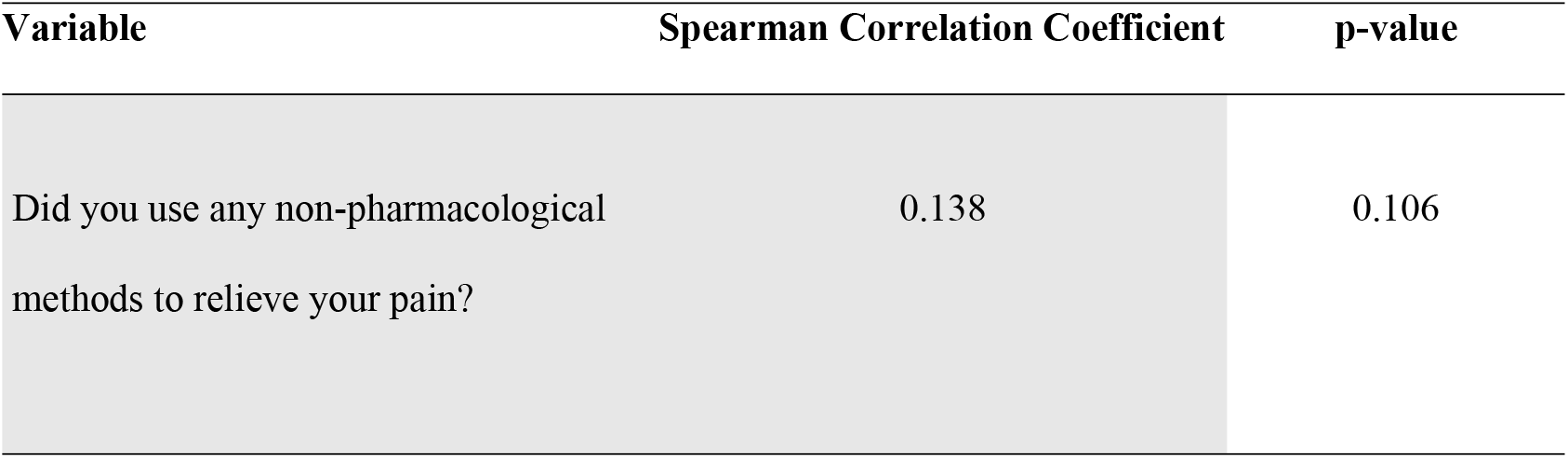
Correlation between the use of NPMs and the wish for more pain treatment.

### 4. Does the use of NPM influence patient satisfaction?

Majority of the participants were highly satisfied 98 (71%) with the overall pain treatment received, with 31 (22.5%) who were highly satisfied. Meanwhile, majority of the patients who were highly satisfied were seen to have used some form of NPMs of pain relief. There was no significant association for satisfaction with pain treatment between the use and non-use of NPMs of pain relief (p-value > 0.05) (Table 7).

**Table 7:** Relationship between the use of NPM and the level of satisfaction of patients with pain treatment.

| Level of satisfaction with pain treatment | Did you use any non-pharmacological methods to relieve your pain |  | Total (n) |
| --- | --- | --- | --- |
|  | Yes (n) | No (n) |  |
| Moderately satisfied | 4 | 5 | 9 |
| Highly satisfied | 55 | 43 | 98 |
| Very highly satisfied | 13 | 18 | 31 |
| Total | 72 | 66 | 138 |

p-value (Cramer's V) = 0.345

A spearman correlation between the use of NPMs of pain relief and over-all satisfaction with pain treatment obtained indicates a significant negative correlation between the type of non-pharmacological treatment and satisfaction with pain treatment (Spearman coefficient = -0.231, p-value = 0.027). Meanwhile, the use of NPMs of pain relief did not have a significant association with over-satisfaction with pain treatment received (p-value > 0.05) (Table 8).

**Table 8:** Satisfaction of patients with pain treatment based on the use of NPMs of pain relief.

| Variable | Spearman Correlation Coefficient | p-value |
| --- | --- | --- |
| Did you use any non-pharmacological methods to relieve your pain? | 0.077 | 0.370 |
| If yes were these non-pharmacological methods helpful in reducing your pain? | -0.231 | 0.027 |

\*. Correlation is significant at the 0.05 level (2-tailed).

## Discussion

The use of NPMs for POP management is intended to minimize the use of analgesics and augment the relief of POP as well as improve the quality of life of patients. In our sub-region, few studies have assessed the use of NPM of POP in caring for patients postoperatively hence the aim of this study.

During the first hours following surgery pain is as its highest level. The initial and most effective method for pain control is to deliver analgesics in the early postoperative period. Again, during the first 48hours postoperative period, patients suffer unavoidable severe to moderate pain so generally combination therapy of opioids and NSAIDs are administered as their postoperative analgesia. [27,25] NPM is often used in POP management after the first 24 hours of surgery [27]

This study revealed that 52.2% of the population representing over half of the population reported the use of NPMs for POP management which could be because health care professionals knew the use of NPMs and so encouraged patients to use them often. This finding is consistent with a study by Koumann et al. where half of their respondents reported the use of NPMs for pain relief [28]

Walking was the most widely used NPM of pain relief accounting for 72.9% of the population in this study. This current study agrees with a study by Ali & Fattah 2020 where they reported walking and deep breathing exercise as the main form of NPMs for managing postoperative pain Even though there have been certain developments in the nurse-to-patient ratio and also doctor-to-patient ratio in Ghana, health care professionals are nonetheless under stress to give out their best health-care services with a particular time frame as they are flooded with a huge number of patients to attend to [30] so the type of NPMs where patients can perform on their own will be recommended for them. This may be why walking was mostly used over the remaining NPMs in this study. Again, walking may be the preferred NPMs used since most low-and middle-income countries of which Ghana is part, lack modern-day equipment in the management of postoperative pain [31].

The male participants were observed to use more NPMs of pain relief as compared to the females in our study which disagrees with other studies where women used more NPMs but with a comparatively insignificant difference. [28].

The majority of the participants representing 51.4% were encouraged by nurses to use NPMs for their pain relief (Table 2) which disagrees with other studies where Yaban 2019, as well as Kidanemariam and colleagues, 2020, reported nurses, have either utilized little or no NPMs in the management of pain [32,33].

The difference in the findings of this current study could be due to adequate training by nurses and other healthcare professionals on the effects of using NPMs for pain relief as has been reported by other studies. [6,34].

Meanwhile, a substantial majority of the patients (51.4%) who used NPMs for their pain relief reported having their pain not relieved which is consistent with reports by previous studies [28]. Whereas a study by Gumus et al., 2020 disagrees with the findings of this current study. [34]. The difference may be because pharmacologic pain treatment was started effectively postoperatively before NPMs were initiated after the first 24 hours or after patients gained pain control.

Moreover, in a systematic review by Ay 2018, it was reported that sociodemographic characteristics which include sex, age, weight, height, duration of anesthesia, duration of surgery, experience, and smoking influence pain severity but are different amongst children and adults. In our current findings, the type of surgery the patients underwent had a statistically significant negative correlation with the use of NPMs for pain relief (Table 4) which is similar to a study by Koumann and friends where they reported that the effectiveness of NPMs is associated to some type of surgeries [27,28].

The literature revealed that health care practitioners’ attitude toward the different types of non-pharmacological pain management makes the efficacious use of these measures questionable [16,6]. In that regard, a higher percentage (90.6%) reported the wish for more pain treatment which is similar to other studies done elsewhere [28].

There was no statistically significant relation to the use of NPMs of pain relief and patients’ desire for more pain treatment (Table 6).

This similarity may be due to more workload on nurses and other healthcare professionals which gives them less time to practice NPMs with the patients and so more attention is given to administering pharmacological treatments.

However, the majority of the participants (71%) were highly satisfied with the overall pain treatment received. Meanwhile, most patients who were highly satisfied were seen to have used some form of NPMs of pain relief. This finding is in contrast to a study done in Nigeria [35].

There was a significant negative correlation between the type of non-pharmacological treatment received and satisfaction with pain treatment (Spearman coefficient = -0.231, p-value = 0.027). However, the use of NPMs for pain relief did not have a significant association with over-satisfaction with pain treatment received (p-value > 0.05) (Table 8)

Patients are generally satisfied with POP management but the use of NPMs should be complimented with pharmacologic treatments to affect overall satisfaction with POP.

## Conclusion

NPMs have been proven to be efficient, inexpensive, and have little or no side effects on patients. It will be appropriate if patients were allowed to have choices as to which NPMs should be used. There should be a need for nurses and other healthcare professionals to educate and offer NPMs to their patients. There should be continuous education on NPMs of postoperative pain to nurses and other healthcare professionals to help promote the efficiency of intervening in postoperative pain with the NPMs.

## Limitations

This study was limited to patients who had had abdominal surgeries hence there have to be caution when relating to other cohorts with other types of surgeries. The study was also done in one tertiary hospital and so caution should be taken when generalizing the findings to a different context.

## Data Availability

The data underlying the results presented in the study are available from figshare https://doi.org/10.6084/m9.figshare.13239449

## Acknowledgments

We would like to express our profound gratitude to all post-operative patients who participated in the study.

## Clinical Trial Registry Name and Registration Number

Not applicable

